# Age-specific changes in mental, physical, lifestyle, and motor function in older adult long-term participants at “kayoinoba” (community gathering places) in a Japanese city: A cohort study

**DOI:** 10.1101/2025.04.23.25326270

**Authors:** Keisuke Kubota, Takaya Abe, Shinichi Shirota, Takashi Nasu, Hiroo Furusawa, Kazuyuki Saotome, Yoshinori Kitabatake, Toyohiro Hamaguchi, Naohiko Kanemura, Toshiyuki Yoshida, Yayoi Amakusa

## Abstract

**Introduction:** This study assessed the mental, physical, and lifestyle functioning of older adult long-term participants (5 years) in a Japanese city’s kayoinoba using the Kihon checklist, 30-second chair stand test (CS-30), and a single leg stance test (SLS test).

**Methods:** This study was conducted as an observational study. Participants were categorized into young-old (<75 years) and old-old (≥75 years) groups based on their initial age.

**Results:** Although total Kihon checklist scores increased significantly at follow-up, no significant difference was found after adjusting for age, suggesting a strong aging effect. Depressed mood (DM) had a relatively pronounced impact on total Kihon checklist scores in the old-old group. The young-old group exhibited a smaller increase in total Kihon checklist scores compared to the old-old group. Motor function remained above that of the general older adults, regardless of age.

**Conclusion:** The findings indicate that long-term participation in kayoinoba, particularly among young-old individuals, improves motor function and social participation frequency, potentially reducing the risk of requiring long-term care.

## Introduction

In Japan, the 2005 revisions to the long-term care insurance system established in 2000 transformed it into a prevention-oriented system, which promoted regional support projects centered on municipal preventive care programs, especially for senior citizens. Additionally, since 2011, various initiatives have been pursued to establish community-based integrated care systems to ensure that older adults can continue living as they would like within regions in which they are accustomed until the end of their lives [1]. According to recent statistics, the older adults ratio of the Japanese population exceeds 28%; moreover, by 2065, about 1 in 2.6 people will be aged ≥65 years, while about 1 in 3.9 people will be aged ≥75 years [2]. Accordingly, there is an urgent need to promote good health, social participation, and employment support for the older adults. To this end, environmental improvements that allow older adult people to voluntarily participate in regional communities have recently attracted attention as a means of coping with Japan’s aging problem.

Preventive care projects in Japan, including long-term care and daily support services, have been developed under the Long-Term Care Insurance (LTCI) framework. These services, such as day care for rehabilitation, daily living support, and visiting care, aim to reduce inactivity risks and help older adults maintain independence [3]. Assessments at the local level allow municipalities to tailor services based on resources and population needs. Within this framework, kayoinoba (community gathering places) has emerged as an effective initiative. It is a community project where older adult leaders form groups in their local areas to engage in physical exercise and social activities [4]. A key feature of kayoinoba is its resident-centered approach, enabling older adults to take active roles in organizing and leading activities. Unlike other preventive care services that rely on professional support, kayoinoba fosters participant contributions, strengthening their connections with others and promoting mutual support within the community. The effectiveness of kayoinoba in terms of preventive care has recently been demonstrated [4–6]. Since 2016, a kayoinoba focused on physical exercise has operated in Koshigaya City, Saitama Prefecture, spanning 41 locations with 851 older adults. A survey of 251 participants assessed their long-term care risk and motor function after six months, revealing that kayoinoba participation was particularly beneficial for those at higher risk of long-term care [7]. These findings underscore the role of kayoinoba in maintaining physical and mental function and reducing long-term care needs.

The long-term sustainability of newly established kayoinoba within communities remains uncertain. In our previous study, only 257 (≈30%) of 851 participants completed the follow-up survey, raising concerns about continued participation and program longevity [7]. Long-term community-based programs offer various benefits, including improved physical and mental health, strengthened social ties, and increased community resilience. Sustained participation enhances social cohesion and supports health equity through collaborative health initiatives [8]. Participatory research has shown significant improvements in empowerment, health behaviors, and community-level changes, particularly for socially disadvantaged populations [9]. However, sustaining long-term programs poses challenges, including declining participation, unengaging content, and limited community support [10]. Additionally, participants’ deteriorating health complicates engagement [11]. Continuous community involvement and adaptive program design are crucial for addressing these challenges and ensuring sustainability [8].

To address these challenges, it is essential to clarify the mental, physical, and functional characteristics of older adults with long-term kayoinoba participation. Demonstrating its benefits could boost engagement and counter declining participation. Additionally, such evidence may enhance local residents’ understanding of kayoinoba, fostering greater support and involvement. Examining its impact on aging-related decline and identifying areas for improvement can validate kayoinoba’s value and guide the development of more sustainable and engaging programs.

Therefore, to facilitate the long-term operation of kayoinoba, this study aimed to survey older adult long-term participants in the kayoinoba of Koshigaya city with respect to their mental, physical, and lifestyle functioning. This study may improve the understanding and participation of local residents, inform new and improved programs, facilitate health maintenance, and revitalize regional exchange.

## Materials and Methods

### Participants

This study employed a mixed study design, analyzing data collected from the kayoinoba preventive care program. Retrospective data were obtained from program records starting in September 2016 (initial evaluation), and prospective data were collected through follow-up evaluations until December 2023 (follow-up). A total of 1132 older adults who had participated in the kayoinoba held at 46 locations in the city were included as study participants. The study objective was orally explained to the participants and written informed consent was obtained. Participants who did not provide informed consent or for whom data could not be collected at either the initial evaluation or follow-up were excluded. This study was designed as an observational study and followed the relevant EQUATOR guideline (STROBE) for observational studies [12]. This study was approved by the ethics review committee of the affiliated institution (approval number: 22037, date: 2022.09.16). The data were accessed for research purposes on 2022.10.01, following ethical approval.

### Activities at the kayoinoba

The kayoinoba program, as previously described in our study [7], incorporates a structured physical exercise component common across all groups. Specifically, the program consists of (i) the performance of dynamic stretches as warm-up in standing position, engaging the whole body, followed by (ii) the “Koshigaya Rakunobi exercise,” which is a set of resistance exercises for the upper and lower limbs using elastic bands, (iii) the “Rakusho exercise,” in which the participants moved between sitting and standing positions as lower limb exercises while singing to improve oral and respiratory function, and (iv) cooling down with full-body static stretches.

Kayoinoba sessions were held once or twice per week, with each session lasting approximately 1.5 to 2 hours. Of this time, 40 minutes to 1 hour was dedicated to physical exercise, while the remaining time was utilized differently across groups. Some groups conducted additional exercises, while others organized tea gatherings or other social activities, reflecting the unique preferences and needs of each community.

### Survey items

Survey items included the Kihon checklist to evaluate mental, physical, and lifestyle function, as well as the 30-second chair stand test (CS-30) to evaluate lower limb strength and endurance, and a single leg stance test (SLS test) to evaluate static balance function. The Kihon checklist is a self-administered questionnaire with 25 yes/no questions regarding living conditions and mental and physical function, with a maximum score of 25 points. A higher Kihon checklist score indicates a higher risk of requiring long-term care. It has been used to screen older adults at risk of requiring long-term care [13–15]. It comprises seven broad domains: instrumental activities of daily living (ADL), physical function (PF), nutritional status (NS), oral function (OF), outdoor activities (OA), cognitive function (CF), and depressed mood (DM). The validity and reliability of the Kihon checklist have been demonstrated in previous studies. It has shown strong correlations with standardized measures of physical function, cognitive function, nutritional state, and depressive mood, validating its use as a comprehensive tool for assessing frailty [15]. Moreover, its predictive validity has been established through longitudinal studies, which demonstrated its ability to predict long-term care dependency and mortality in community-dwelling older adult populations [14]. These findings confirm the checklist’s utility as a screening tool for identifying individuals at risk of requiring support or care.

The SLS test measures how long one can lift one leg and continue standing on the other leg with one’s eyes open [16]. A study has demonstrated that the one-leg standing time test has high test-retest reliability in older adult populations, with consistent results across repeated measures. Moreover, its validity has been supported by its strong correlation with functional balance and mobility assessments, indicating its effectiveness as a screening tool for physical frailty and fall risk in older adults [17]. CS-30 measures the number of times one can stand up from a chair in 30 seconds [18,19]. This test has been shown to have high reliability and validity, making it a robust tool for assessing lower extremity strength and physical performance in older adult populations. Its utility as a screening tool for physical frailty and sarcopenia has also been widely recognized [20].

The aforementioned survey items were obtained at the “initial” evaluation upon participation in the kayoinoba and the "follow-up" evaluation conducted after 5 years. These data were collected and managed by the Koshigaya Rehabilitation Liaison Council.

### Statistical analysis

To evaluate changes in mental, physical, and lifestyle functioning, total Kihon checklist scores were analyzed at initial and follow-up evaluations. Statistical analyses included:

1, Data Normality Testing:

- Kolmogorov-Smirnov test for normality.
- Normally distributed data were analyzed using a t-test, while non-normally distributed data were assessed with the Wilcoxon signed-rank test.
2, Within-Group Comparisons (Adjusted for Age):

- T-test for normally distributed data; Wilcoxon signed-rank test for non-normal data.
- Changes between initial and follow-up evaluations were analyzed as a single group.
- ANCOVA for normally distributed variables (e.g., total Kihon checklist scores, SLS test, and CS-30) with age as a covariate.
- Quade test for non-normal variables.
3, Within-Group Comparisons by Age Group (Adjusted for Age):

- Participants were classified as <75 years (young-old) or ≥75 years (old-old).
- ANCOVA for normally distributed data; Quade test for non-normal data.
4, Contribution Ratio and Correlation Analysis:
To evaluate the impact of changes in each sub-item on the total Kihon checklist scores, the following steps were performed:

- Initial and follow-up scores for each sub-item were standardized using their respective maximum values.
- Changes in standardized and total scores (Δtotal) were computed.
- Contribution ratios were calculated as the proportion of sub-item scores relative to total score changes, with small total score changes adjusted to prevent extreme ratios.
- Pearson’s correlation was used to assess the relationship between sub-item and total score changes.
- Sub-items with high contribution ratios and strong correlations significantly influenced total Kihon checklist scores.
5, Age and Kihon Checklist Score Variation:

- Pearson’s correlation examined age at follow-up and total Kihon checklist score variation.
- A high positive correlation would suggest that older individuals experience greater declines in total Kihon checklist scores, reflecting worsened mental, physical, and lifestyle functioning.

All statistical analyses were performed using MATLAB 2024a, and the significance level was set at p = 0.05. No multiple comparison corrections (e.g., Bonferroni correction) were applied in this study, as the analyses were exploratory in nature, and the results were not intended to independently confirm multiple hypotheses.

## Results

A detailed flowchart outlining participant selection and exclusion is presented in S1 Fig. Of the 1132 study participants, 615 attended only the initial evaluation, 208 participated only in the follow-up assessment, and 186 were excluded due to data irregularities, totaling 1009 exclusions. Consequently, 123 older adults who had participated in kayoinoba for five years were included in the final analysis.

S1 Table shows the comparison between initial evaluation and follow-up. Significant differences were observed, with age and total Kihon checklist scores being significantly higher and SLS test results significantly lower at follow-up compared to the initial evaluation, while no significant difference was found for CS-30. A Quade test with adjustment for age revealed that age was significantly higher, while SLS test and CS-30 results were significantly lower at follow-up. No significant difference was found for the total Kihon checklist score.

Table 1 shows the comparison between the initial evaluation and follow-up in the young-old group. Significant differences were observed, with age and total Kihon checklist scores being significantly higher and SLS test results significantly lower at follow-up, while no significant difference was found for CS-30. However, after adjusting for age, none of these variables showed significant differences. Table 2 presents the comparison between the initial evaluation and follow-up in the old-old group. Significant differences were observed, with age and total Kihon checklist scores being significantly higher at follow-up, while no significant differences were found for SLS test and CS-30 results. After adjusting for age, SLS test and CS-30 results were significantly lower at follow-up, but no significant difference was found for total Kihon checklist scores.

**Table 1.** Comparison between initial evaluation and follow-up in the young-old group (N = 60)

| Young - old group | Initial<br>evaluation<br>(±SD) | Follow-up<br>(±SD) | 95% CI | Effect<br>size | P-values | After adjustment<br>for P-values<br>in age |
| --- | --- | --- | --- | --- | --- | --- |
| Age | 70.23 (±4.54) | 74.96 (±4.72) | 2.21 to 7.60 | 3.27 | < 0.001 | |
| Total Kihon checklist<br>scores (point) | 3.37 (±2.69) | 4.05 (±2.94) | 0.04 to 0.58 | 0.29 | 0.03 | 0.16 |
| SLS test (sec) | 48.33 (±43.88) | 31.09 (±24.39) | -0.77 to -0.26 | -0.49 | < 0.001 | 0.67 |
| CS-30 (time) | 22.15 (±5.11) | 21.15 (±7.60) | -0.35 to 0.11 | -0.11 | 0.39 | 0.13 |

Abbreviations: SD, standard deviation; CI, confidence interval; SLS test, single leg
stance test; CS-30, 30-second chair stand test

**Table 2.** Comparison between initial evaluation and follow-up in the old-old group (N = 63)

| Old - old group | Initial<br>evaluation<br>(±SD) | Follow-up<br>(±SD) | 95% CI | Effect<br>size | P-values | After adjustment<br>for P-values<br>in age |
| --- | --- | --- | --- | --- | --- | --- |
| Age | 79.49 (±2.62) | 83.69 (±2.70) | 3.42 to 5.68 | 4.20 | < 0.001 | |
| Total Kihon checklist<br>scores (point) | 4.02 (±3.06) | 5.81 (±3.76) | 0.30 to 0.84 | 0.60 | < 0.001 | 0.99 |
| SLS test (sec) | 25.83 (±36.43) | 20.74 (±22.26) | -0.40 to 0.10 | -0.18 | 0.19 | <0.001 |
| CS-30 (time) | 19.42 (±5.78) | 18.06 (±8.76) | -0.32 to 0.09 | -0.15 | 0.26 | 0.03 |

Abbreviations: SD, standard deviation; CI, confidence interval; SLS test, single leg
stance test; CS-30, 30-second chair stand test

Regarding the contribution ratio of each sub-item to the change in total Kihon checklist scores and the correlation analysis (S2 Table), items for DM showed the highest contribution ratio and correlation coefficient. This was followed by PF and ADL for contribution ratios and ADL and PF for correlation coefficients. In the young-old group, contribution ratios and correlation coefficients tended to be higher for PF and CF. In the old-old group, contribution ratios and correlation coefficients tended to be higher for DM (S3 Table).

There was a significant positive correlation between the range of changes in total Kihon checklist scores and age at follow-up (S2 Fig).

Table 3 presents the between-group comparisons of total Kihon checklist scores between the young-old and old-old groups at both the initial evaluation and follow-up.

**Table 3.** Comparison of total Kihon checklist scores by group at initial evaluation and follow-up (Young-old group: N = 60, Old-old group: N = 63)

| | Young-old group (±SD) | Old-old group(±SD) | 95% CI | Effect size | P-values |
| --- | --- | --- | --- | --- | --- |
| At initial evaluation<br>total Kihon checklist score | 3.37 (±2.69) | 4.02 (±3.06) | -0.13 to 0.50 | 0.23 | 0.33 |
| At follow-up<br>total Kihon checklist score | 4.05 (±2.94) | 5.81 (±3.76) | 0.16 - 0.88 | 0.52 | 0.005 |

Abbreviations: SD, standard deviation; CI, confidence interval

No significant differences were observed at the initial evaluation, while at follow-up, the old-old group had significantly higher total Kihon checklist scores than the young-old group.

## Discussion

We observed a significant increase in total Kihon checklist scores at follow-up. However, as no significant difference was found after adjustment for age (S1 Table), the observed changes after 5–6 years might be strongly influenced by aging. Notably, DM showed the highest correlation coefficients and contribution ratios to the total Kihon checklist scores (S2 Table). A systematic review and meta-analysis reported prevalence rates of 7.2% for major depressive disorder and 17.1% for depressive disorders in older adults aged >75 years [21,22]. This further supports the idea that DM contributes to the aging-related increase in total Kihon checklist scores. The amount of exercise required to effectively improve depressive symptoms is ≥320 Metabolic Equivalent for Tasks-min per week [23]. Although we could not determine the daily amount of exercise among our participants, it may have been insufficient to prevent depressive tendencies. Similar to exercise, preventing social isolation has been shown to effectively protect against depressive tendencies [24]. Participating in kayoinoba has been shown to increase the frequency of social participation in daily life [6]. Accordingly, even for the kayoinoba in Koshigaya city, it may be beneficial to add activities other than physical exercise, such as communication, and encourage social participation activities outside of kayoinoba. However, since depression cannot be determined only using a Kihon checklist, our findings merely suggest an approach for preventing depressive tendencies.

We observed within-group differences in the change in total Kihon checklist scores between the initial evaluation and follow-up for each group (young-old and old-old groups) (Tables 1 and 2). First, there was no significant within-group difference between the initial evaluation and follow-up in either group after adjustment for age, which could be similarly attributed to the strong effect of aging. However, there were between-group differences in the contribution ratios and correlation coefficients. In the young-old group, PF and CF tended to show high contribution ratios and correlation coefficients (S3 Table). Both motor and cognitive function decline with age [25,26]. Additionally, physical activity has been positively correlated with cognitive function [27]. Accordingly, these two items have an inevitable effect on total Kihon checklist scores. However, in relative terms, each sub-item showed a comparatively uniform distribution. Therefore, in the young-old group, all sub-items, including PF and CF, showed a comparable effect on total Kihon checklist scores. However, in the old-old group, DM (depression) tended to have a relatively pronounced effect compared with other items (S3 Table). The mean age of participants in this group at follow-up was 83.7 years, which is close to the reported peak incidence of depression at 85–89 years of age [28]. This finding may explain the prominent impact of DM observed in this study. Research has demonstrated that depression in older adults is closely linked to physical frailty and cognitive decline, forming a cycle where depression exacerbates both conditions [29]. This relationship is particularly pronounced in the old-old group, who are more vulnerable to physical and psychological stressors due to advanced age and reduced coping resources, a phenomenon described as “psychosocial frailty” [29]. Furthermore, inadequate social support, including diminished social networks and the loss of close relationships such as spouses or peers, significantly contributes to depression in late life [30]. This lack of social support not only exacerbates depressive symptoms but also negatively affects physical and cognitive health, with a particularly strong impact observed in the old-old population. These findings underscore the importance of addressing depression as a critical factor in preventing overall functional decline in the old-old group.

Additionally, the total Kihon checklist showed no significant difference between the young-old and old-old groups at the initial evaluation. However, at the follow-up evaluation, the total Kihon checklist scores were significantly higher in the old-old group than in the young-old group (Table 3). This suggests a larger increase in total Kihon checklist scores in old-old than in young-old people. Furthermore, we observed a significant positive correlation between age at follow-up and the change in total Kihon checklist scores (S2 Fig). These findings align with prior research indicating that functional decline accelerates with age and is more pronounced in the old-old population than in the young-old population. Studies have shown that the old-old experience steeper declines in both physical and cognitive functions. For instance, Rönnlund et al. [31] reported that cognitive abilities such as processing speed and verbal memory decline more sharply in the old-old group. Similarly, Harada et al. [32] highlighted that physical frailty, including reduced balance and muscle strength, becomes increasingly prevalent with advancing age. These age-related declines, particularly in the old-old group, contribute to the deterioration of total Kihon Checklist scores, reflecting greater risks of functional disabilities and the need for long-term care. This highlights the importance of addressing these declines proactively, especially among young-old individuals. Early participation in activities such as exercise and social exchange may help mitigate the progression of physical and cognitive decline, thereby reducing the risk of requiring long-term care. These findings emphasize the value of both prompt and sustained engagement in kayoinoba among older adults.

Regarding CS-30, the young-old group showed no significant within-group differences between the initial evaluation and follow-up, while the old-old group showed significant within-group differences after adjustment by age (Tables 1 and 2). This suggests that the CS-30 is relatively stable in the young-old group, regardless of age. The CS-30 is a highly reliable method for screening the risk of sarcopenia, with cutoff values of 15 and 17 times for women and men, respectively [20]. For the young-old group, the average CS-30 values were 22.2 and 21.2 times at initial and follow-up evaluations, respectively. This demonstrated that CS-30 for young-old people may be maintained even over the long term and was higher than the national average. Age showed a significant effect in the old-old group, but the average values at the initial and follow-up evaluations were 19.4 and 18.1 times, respectively, both above the cutoff value. Therefore, even though there was a decrease in CS-30 in the old-old group, they still presented better physical function than older adults in the general population. The decline in CS-30 performance in the old-old group may not be solely due to muscle weakness but also cognitive decline. As CS-30 involves repetitive sit-to-stand movements, executive function and balance coordination play a role in performance [33]. Additionally, psychological factors such as depressive tendencies, which were strongly correlated with total Kihon checklist scores in the old-old group, may have contributed to reduced performance [22]. These findings suggest that maintaining CS-30 function requires not only physical training but also cognitive and mental health interventions.

The SLS test showed no significant difference after adjustment for age in the young-old group. In the old-old group, although no significant differences were observed in Wilcoxon’s signed-rank test, differences were observed after adjusting for age (Tables 1 and 2). Similar to CS-30, this demonstrated an effect of age on the SLS test in the old-old, but not young-old group. The SLS test is part of the Berg balance scale, which is an index for screening lower limb muscle strength and balance function [34]. Among older adults, the average SLS test result is 15.1 s for women [35]. In this survey, the average SLS test results among the young-old group at the initial and follow-up evaluations were 48.3 s and 31.1 s, respectively. In the old-old group, the average SLS test results at the initial and follow-up evaluations were 25.8 s and 20.7 s, respectively. In both groups, the measured values remained above the general average. Therefore, the program relatively maintained motor function. The decline in SLS performance in the old-old group may be influenced not only by muscle weakness but also by cognitive and neurological factors. Balance control involves sensory integration and motor coordination, both of which deteriorate with age [32]. Additionally, attention and executive function are crucial for maintaining postural stability, and recent systematic reviews have shown that executive function, processing speed, and global cognition exhibit significant associations with balance performance in older adults [36]. These findings suggest that to maintain balance and prevent falls, interventions should incorporate not only physical training but also cognitive and sensory-motor exercises.

To further enhance the social and mental health benefits of kayoinoba, incorporating non-physical activities is a promising approach. While the current program has demonstrated effectiveness in maintaining motor functions and supporting social participation, additional activities targeting mental and emotional well-being could significantly broaden its impact. Specific communication-based activities, such as group discussions, storytelling sessions, and peer-led workshops, can foster interpersonal connections. Similarly, cognitive stimulation activities like memory games, puzzles, or creative art workshops may help improve mental well-being. Implementing these activities, however, poses challenges such as securing facilitators with the necessary skills, providing appropriate venues, and ensuring consistent participation. Moreover, tailoring these activities to meet the needs of young-old and old-old participants is essential to maximize effectiveness. For the young-old group, activities emphasizing active social exchange and skill-building may be more engaging, while for the old-old group, activities focused on gentle cognitive stimulation and emotional support may be more suitable. By adopting such tailored approaches, kayoinoba programs can better address the diverse needs of their participants and enhance their overall impact on social and mental health outcomes.

### Limitations

The present study had some limitations. Firstly, although we obtained relatively favorable results regarding motor function using the SLS test and CS-30 tests, we found that the sub-item of PF in the Kihon checklist was correlated with an increase in total Kihon checklist scores. This could be attributed to the fact that the SLS test and CS-30 are comprehensive motor function tests that can be performed objectively, while PF based on the Kihon checklist sub-items involves subjective questions regarding physical function in daily life. It may be important to improve motor-related Kihon checklist sub-items to better reflect daily life. Secondly, while a power analysis indicated that a sample size of 128 participants would be required to achieve a power of 0.8 (80%) with an alpha level of 0.05 and an effect size of 0.25, the study included 123 participants who met the inclusion criteria. Although the sample size was slightly smaller than the calculated requirement, the findings provide meaningful insights into the target population. Future studies could aim to recruit a larger sample size to further validate and generalize the findings. Thirdly, this study acknowledges that depression cannot be diagnosed using the Kihon checklist. While the checklist includes items related to mood, it is primarily designed to evaluate comprehensive long-term care risks rather than to screen for depressive tendencies. Consequently, there is potential for misclassification or underestimation of depressive symptoms, as the checklist does not comprehensively evaluate the psychological or emotional dimensions of depression. Integrating other validated depression screening tools, such as the Geriatric Depression Scale (GDS), could provide a more accurate and detailed understanding of depressive symptoms among participants. This would enhance the findings and contribute to a more comprehensive evaluation of mental health in this population. Finally, the present study could not strictly determine whether the observed improvements were directly attributable to kayoinoba or influenced by other unmeasured factors, such as participant motivation or community support. However, we emphasize that kayoinoba is beneficial as a comprehensive initiative, including such factors. Based on the findings of this study, future research should explore the effectiveness of kayoinoba through experimental designs, such as modifying the contents of the program, to establish its causality and further evaluate its benefits.

## Conclusions

Our findings demonstrated a significant increase in total Kihon checklist scores at follow-up. However, there was no significant difference after adjustment for age, which suggests a strong effect of aging. In the old-old group, depressed mood (DM) tended to have a relatively pronounced effect on total Kihon checklist scores compared with other sub-items. Considering that the exercise regimen within the program appeared to be insufficient for preventing depressive tendencies, it is important to promote activities involving communication and social participation in such a program. Moreover, it was suggested that the increase in total Kihon checklist scores was smaller in the young-old group than in the old-old group. This suggests that long-term participation in kayoinoba, especially among young-old individuals, can increase motor function and the frequency of social participation, thereby facilitating the prevention of the risk of requiring long- term care.

## Data Availability

All relevant data are within the manuscript and its Supporting Information files.

## Acknowledgments

The authors thank all participants and gratefully acknowledge the assistance and support provided by the Koshigaya Rehabilitation Liaison Council.

## Supporting information

**S1 Figure. Flowchart showing the selection of participants for analysis**

**S2 Figure. Correlation between changes in total Kihon checklist scores and age at the time of follow-up (N = 123)**

**S1 Table. Comparison between initial evaluation and follow-up (N = 123)**

**S2 Table. Contribution ratio and correlation coefficient of each item for changes in the total Kihon checklist scores (N = 123)**

**S3 Table. Contribution ratio and correlation coefficient of each item for changes in the total Kihon checklist scores (Young-old group: N = 60, Old-old group: N = 63)**

